# A differential regulatory T cell signature distinguishes the immune landscape of COVID-19 hospitalized patients from those hospitalized with other respiratory viral infections

**DOI:** 10.1101/2021.03.25.21254376

**Authors:** Sarah C. Vick, Marie Frutoso, Florian Mair, Andrew J. Konecny, Evan Greene, Caitlin R. Wolf, Jennifer K. Logue, Jim Boonyaratanakornkit, Raphael Gottardo, Joshua T. Schiffer, Helen Y. Chu, Martin Prlic, Jennifer M. Lund

## Abstract

SARS-CoV-2 infection has caused a lasting global pandemic costing millions of lives and untold additional costs. Understanding the immune response to SARS-CoV-2 has been one of the main challenges in the past year in order to decipher mechanisms of host responses and interpret disease pathogenesis. Comparatively little is known in regard to how the immune response against SARS-CoV-2 differs from other respiratory infections. In our study, we compare the peripheral blood immune signature from SARS-CoV-2 infected patients to patients hospitalized pre-pandemic with Influenza Virus or Respiratory Syncytial Virus (RSV). Our in-depth profiling indicates that the immune landscape in patients infected by SARS-CoV-2 is largely similar to patients hospitalized with Flu or RSV. Similarly, serum cytokine and chemokine expression patterns were largely overlapping. Unique to patients infected with SARS-CoV-2 who had the most critical clinical disease state were changes in the regulatory T cell (Treg) compartment. A Treg signature including increased frequency, activation status, and migration markers was correlated with the severity of COVID-19 disease. These findings are particularly relevant as Tregs are being discussed as a therapy to combat the severe inflammation seen in COVID-19 patients. Likewise, having defined the overlapping immune landscapes in SARS-CoV-2, existing knowledge of Flu and RSV infections could be leveraged to identify common treatment strategies.

**Highlights:**

1. The immune landscapes of hospitalized pre-pandemic RSV and influenza patients are similar to SARS-CoV-2 patients
2. Serum cytokine and chemokine expression patterns are largely similar between patients hospitalized with respiratory virus infections, including SARS-CoV-2, versus healthy donors
3. SARS-CoV-2 patients with the most critical disease displayed unique changes in the Treg compartment
4. advances in understanding and treating SARS-CoV-2 could be leveraged for other common respiratory infections

**Graphical Abstract:** 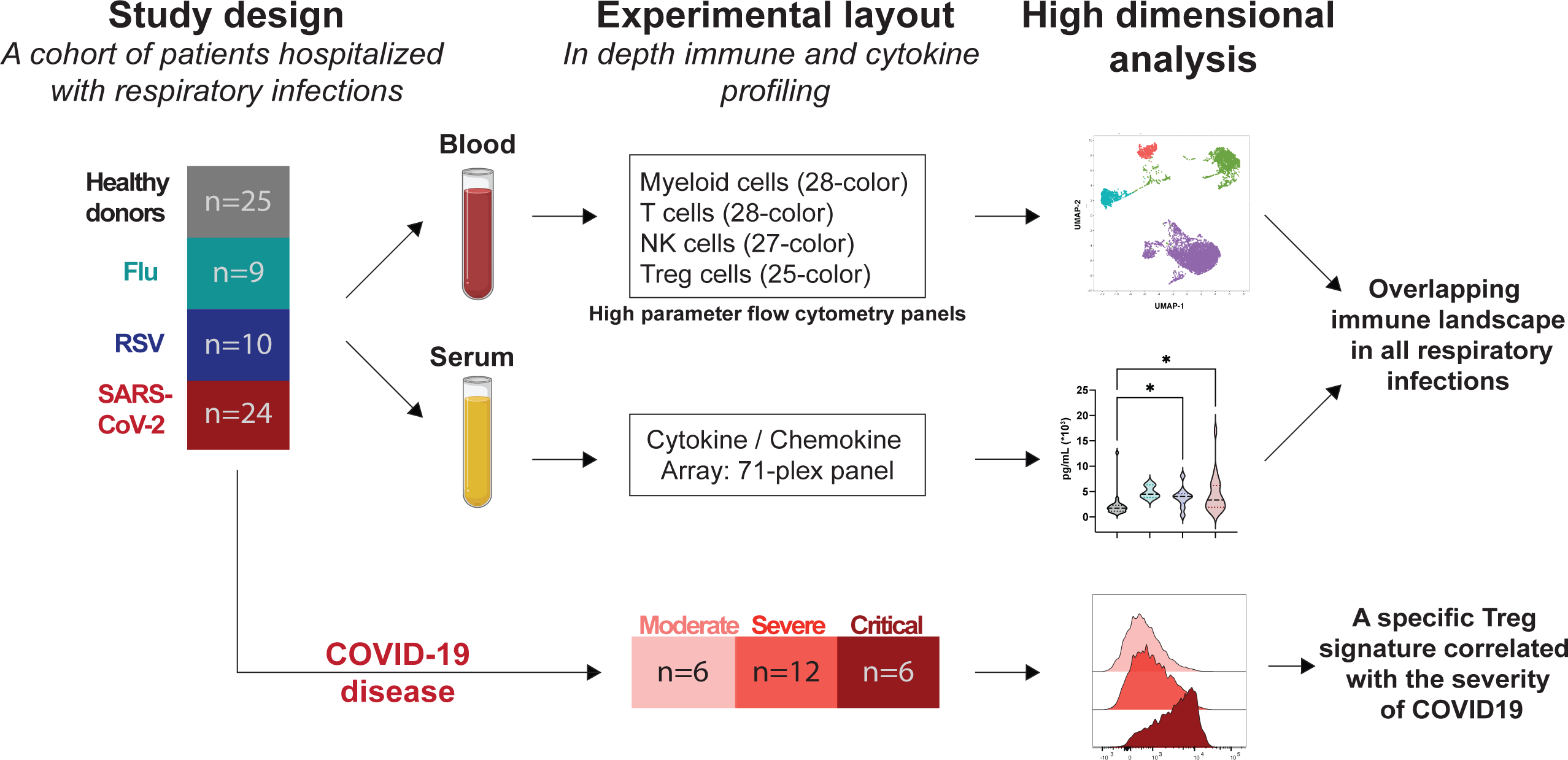

## Introduction

The current coronavirus (CoV) pandemic began in Wuhan, China in 2019 with an outbreak of what would later be designated SARS-CoV-2^1–3^. To date, SARS-CoV-2 has led to devastating disease (called Coronavirus Disease 2019 (COVID-19)), death, and economic instability on a global scale^4^. Despite the unprecedented rapid design and large-scale testing of SARS-CoV-2 vaccines, vaccine supply shortages, vaccine hesitancy, delays in global implementation, and emerging variants raise concerns that SARS-CoV-2, as well as the next pandemic respiratory infection, will continue to pose a threat to humans, underscoring the vital need for identification of additional therapeutics.

Many investigations to date have focused on characterizing the immune responses to natural SARS-CoV-2 infection in an effort to understand disease pathogenesis and reveal potential therapeutic targets. We reasoned that these extraordinary efforts to understand COVID-19 disease pathogenesis and improve treatment options could be leveraged for other respiratory infections if there is substantial congruence in the underlying immune response. In the vast majority of such SARS-CoV-2 studies, immune responses have been measured using human blood samples, comparing healthy controls to asymptomatic patients with SARS-CoV-2 or COVID-19 patients with varying degrees of disease severity^5^. Both the innate and adaptive arms of the immune response to SARS-CoV-2 have been profiled to date. Initial studies focusing on innate immunity demonstrated that the type I and type III interferon (IFN) response is blunted in early stages of the response to SARS-CoV-2, though IL-6 and chemokines are elevated ^6, 7^. Notably, this varied from the response to other respiratory viral infections, including human parainfluenza virus 3 and respiratory syncytial virus (RSV), which induced potent type I and III IFN responses^6^ and could suggest fundamental differences in the immune landscape of different respiratory infections. Further, data derived from comparing healthy controls and patients with severe COVID-19 identified an early reduction in type I IFNs in patients with the most severe or critical disease, as well as enhanced pro-inflammatory IL-6 and TNF responses^8–12^. In association with this depressed type I IFN response, patients with critical cases of COVID-19 have a corresponding decrease in frequency of professional type I IFN producing cells, plasmacytoid dendritic cells (pDC)^8^. The frequency of NK cells was significantly diminished in SARS-CoV-2 patients with acute respiratory distress syndrome (ARDS) as compared to healthy donors, though patients with more severe disease had NK cells with increased expression of activation and cytotoxic molecules^13, 14^. Additionally, increased frequencies of neutrophils have been identified in patients with severe COVID-19 as compared to patients with more mild disease or healthy donors^10, 11, 13, 15^, congruent with a hyper-inflammatory state.

Patients infected with SARS-CoV-2 also raise detectable adaptive immune responses, in the form of both B and T cell responses specific to SARS-CoV-2^16–24^. Additionally, circulating conventional T cell phenotypes have been extensively profiled in patients infected with SARS-CoV-2 with varying degrees of disease, from asymptomatic or mild to critical disease, and several differences in the dynamics of immune cells have been noted, including increased abundance of activated T cells in patients with the most severe COVID-19 disease as compared to healthy controls^10, 13, 23–25^. Moreover, two groups have noted a significant decrease in abundance of circulating regulatory T cells (CD3+CD4+CD25+CD127lo) in patients with severe COVID-19 as compared to patients with non-severe disease or healthy donors^11, 26^. Notably, one study identified a decrease in airway regulatory T cells in patients with COVID-19 compared to healthy controls ^27^, raising the possibility that a Treg deficit at the lung site could be contributing to disease.

Altogether, these data suggest that a dysregulated state of hyper-inflammation is associated with severe COVID-19. However, it remains largely unclear if this dysregulated hyper-inflammatory state is unique to COVID-19 or is a feature of severe disease with respiratory viral infections more generally. A recent study comparing inflammatory profiles in patients infected with SARS-CoV-2 or influenza virus (Flu) found several notable differences between such patients. These consisted of lower cytokine levels and reduced circulating monocyte counts in patients with SARS-CoV-2 as compared to Flu, although circulating lymphocyte counts did not differ in patients with the two distinct infections^28^. They concluded that SARS-CoV-2 patients have a less inflamed peripheral immunotype as compared to patients infected with Flu, though other respiratory viruses were not examined. Thus, we designed a study wherein circulating immune signatures were compared among healthy human donors and hospitalized patients with SARS-CoV-2, Flu, or respiratory syncytial virus (RSV) infection. Hospitalized patients infected with either of the three viruses were further classified as having moderate, severe, or critical disease based on the type of provided oxygen supplementation, thereby allowing for comprehensive comparisons of immune cell abundance and phenotype across a range of disease severity. In general, our deep immune profiling revealed similar cellular and cytokine immune landscapes in hospitalized patients infected with SARS-CoV-2, Flu or RSV compared to healthy donors. However, unique to COVID-19 patients with the most critical disease was a significant increase in the frequency of regulatory T cells (Treg) in the circulation, as well as phenotypic changes indicating increased suppressive capacity and tissue-migratory patterns. Our novel findings have clinical implications, as treatments used for COVID-19 may be useful in mitigating severe Flu or RSV, as well as future pandemic respiratory diseases. Furthermore, Tregs may provide a potential therapeutic target for COVID-19.

## Results

### Deep immune profiling of a unique cohort of patients reveals shared circulating immune cell composition between respiratory infections

We analyzed PBMCs from a unique cohort of age-and sex-matched patients hospitalized with respiratory infections including Flu, RSV, or SARS-CoV-2 compared to PBMCs from healthy donors (**Figure 1A**). The patients infected with SARS-CoV-2 required varying degrees of oxygen supplementation, and patients experienced varying COVID-19 outcomes, from moderate disease to death (**Table 1**). The PBMCs from hospitalized patients with RSV or Flu A or B were all collected before the SARS-CoV-2 pandemic. To extensively characterize the cellular immunotypes present in the peripheral blood of patients hospitalized with Flu, RSV, or SARS-CoV-2 infection compared to healthy donors, we combined several high-parameter flow cytometry panels to profile myeloid cells, T cells, NK cells, or regulatory T cells (Treg) (**Supplemental Table 1**). For exploratory analysis of this high-dimensional data set we utilized clustering by Flow-SOM^29, 30^ and dimensionality reduction with uniform manifold approximation projection (UMAP)^31^, which revealed strikingly similar distributions of cell populations between healthy donors and all three respiratory infections (**Figure 1B**). A heatmap of markers to identify cell populations distinguished the main clusters as Lin^-^ HLADR^+^ myeloid cells, B cells, T cells, and NK cells (**Figure 1C**). A key feature of SARS-CoV-2 infection that has emerged through recent studies is lymphopenia^2, 7, 32, 33^.

**Figure 1.**
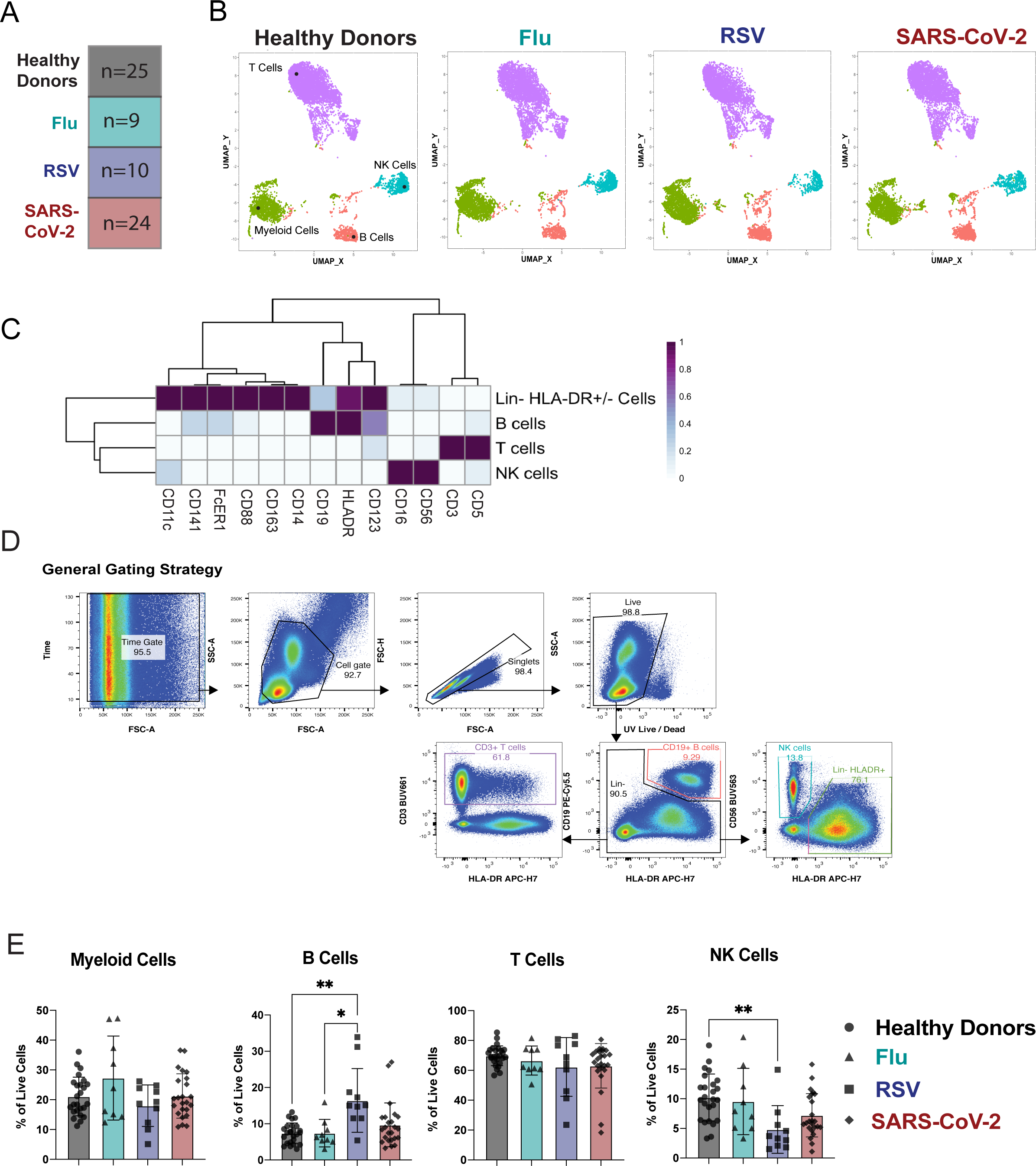
Deep immunological profiling of a unique cohort of patients reveals circulating immune profiles between respiratory infections. A) Overview of the cohort. The number in each box indicates the number of donors per group. Criteria of inclusion are depicted in Table 1. Among the SARS-CoV-2 patients, 4 died from COVID-19. B-D) Previously frozen PBMCs isolated from each group of the cohort were stained using the APC panel (See Supplemental Table 1). B) FlowSOM was used to visualize the main immune cell populations found in the PBMCs from healthy donors or infected patients. C) Heatmaps generated by FlowSOM and used to identify the main immune population. (D-E) (D) Manual gating used to assess the frequencies of the main immune subsets, (E) bar graphs showing these frequencies for each group of our cohort. All data include at least 9 patients per group (Table 1) and are represented as mean ± SD. Statistical analyses were performed using Kruskal-Wallis test. * P < 0.05; ** P < 0.01.

**Table 1.** Demographic and clinical information for study patient cohorts.

|  | Healthy Donors | Flu | RSV | SARS-CoV-2 |
| --- | --- | --- | --- | --- |
| **Number of Patients** | 25 | 9 | 10 | 24 |
| **Age median, (range)** | 60, (33-79) | 59, (36-70) | 57.5, (38-71) | 62, (23-88) |
| **Sex (female/male)** | (12/13) | (4/5) | (4/6) | (11/13) |
| **Race n, (%)** |  |  |  |  |
| White | 20 (80) | 4 (44.4) | 5 (50) | 11 (45.9) |
| Black/African American | 1 (4) | 2 (22.3) | 3 (30) | 3 (12.5) |
| Native Hawaiian/Pacific Islander |  | 1 (11) |  | 1 (4.1) |
| American-Indian/Alaskan Native |  |  | 2 (20) |  |
| Hispanic | 1 (4) | 2 (22.3) |  | 6 (25) |
| Asian | 3 (12) |  |  | 3 (12.5) |
| **Days since symptom onset to sample collection median, (range)** | *n.a.* | 7+, (0-7+)* | 14, (2-21) | 14.5, (2-47) |
| **Comorbidities n** |  |  |  |  |
| None | 17 | 3 | 2 | 7 |
| Diabetes mellitus | 1 | 1 |  | 9 |
| Hypertension | 3 |  |  | 12 |
| Chronic liver disease |  |  | 4 | 2 |
| Chronic kidney disease |  |  | 2 | 5 |
| Congestive heart failure |  | 1 | 2 | 6 |
| Cardiovascular disease |  | 3 | 3 | 3 |
| Asthma | 3 | 3 | 2 | 2 |
| COPD/emphysema | 1 | 1 | 3 |  |
| Other chronic lung disease |  |  |  | 3 |
| Sleep apnea |  |  |  | 7 |
| Malignancy |  |  | 2 | 3 |
| **ACTT Clinical Status Categories n** | *n.a.* |  |  |  |
| Not requiring supplemental O2 – ongoing medical care |  | 4 | 6 | 6 |
| Required supplemental O2 |  | 3 | 0 | 8 |
| Non-invasive ventilation or high flow O2 device |  | 1 | 1 | 4 |
| Intubation, ECMO |  | 1 | 3 | 2 |
| Death |  | 0 | 0 | 4 |
| **Outcome n, (%)** | *n.a.* |  |  |  |
| discharged |  | 9, (100) | 10, (100) | 20, (83.3) |
| death |  | 0 (0) | 0 (0) | 4, (16.7) |

Lymphopenia can also result from other respiratory infections such as Flu and RSV, but this generally occurs early after onset of symptoms and is rapidly resolved^34^. Thus, we wanted to compare alterations in immune subsets across respiratory infections to determine if patients within our SARS-CoV-2 cohort were experiencing similar levels of immune alteration to patients with RSV or Flu versus healthy donors. Manual gating of the flow cytometry data by a conventional gating strategy to determine the main immune populations confirmed the observations seen in the meta-clustering data from FlowSOM (**Figure 1D**). In the Lin^-^HLA-DR^+^ compartment, we did not see significant alterations in the overall frequency across groups. We observed a significant increase in the frequency of B cells in patients hospitalized with RSV compared to patients hospitalized with Flu or to healthy donors. We saw no significant alterations in the frequency of total circulating T cells across all respiratory infections compared to healthy donors. Finally, the frequency of NK cells was significantly reduced only in patients infected with RSV compared to healthy donors (**Figure 1E**). While previous studies have indicated a reduction in NK cell populations after infection with SARS-CoV-2 as compared to healthy donors^13, 14^, our data indicates that this phenomenon is likely not specific to SARS-CoV-2 infection but is seen across additional respiratory infections as well. While the frequencies of these cell population may not directly correlate with the total number of cells found in the blood, we concluded that the overall immune populations remain similar between respiratory infections.

### A decreased frequency of dendritic cell subsets is common across hospitalized individuals with respiratory infections compared to healthy donors

For an in-depth analysis of these immune cell populations, we leveraged 4 high-parameter flow cytometry panels focusing on antigen presenting cells (APC panel), NK cells (NK cell panel), as well as T cells (T cell panel) and Tregs (Treg panel) (**Supplemental Table 1**). For APC, we followed recently suggested phenotyping guidelines to separate classical and non-classical monocytes, as well as 4 distinct dendritic cell (DC) subsets: the pDC, cDC1, cDC2 and the newly defined inflammatory cDC3^35, 36^ (**Supplemental Figure 1A**). None of the infections led to a significant alteration in frequency of classical monocytes compared to healthy donors. While a decrease in non-classical monocyte frequency has been reported with SARS-CoV-2 infection^37^, our data only showed a significant decrease of the non-classical monocytes for Flu while a trend toward reduction was observed for RSV and SARS-CoV-2 infected patients (**Figure 2A**). In a similar manner as Zhou *et al*., we observed a reduced frequency across several DC subsets in SARS-CoV-2 compared to healthy donors^38^. Of note, this decrease in the frequency of pDC, cDC1, cDC2, and inflammatory cDC3 was also observed in Flu and RSV-infected patients (**Figure 2B**). When we assessed the frequencies of the CD56^bright^CD16^-^ as well as the CD56^dim^CD16^+^ NK cell subsets (**Supplemental Figure 1B**), we found no significant differences when compared across respiratory infections and healthy donors (**Figure 2C**).

**Figure 2.**
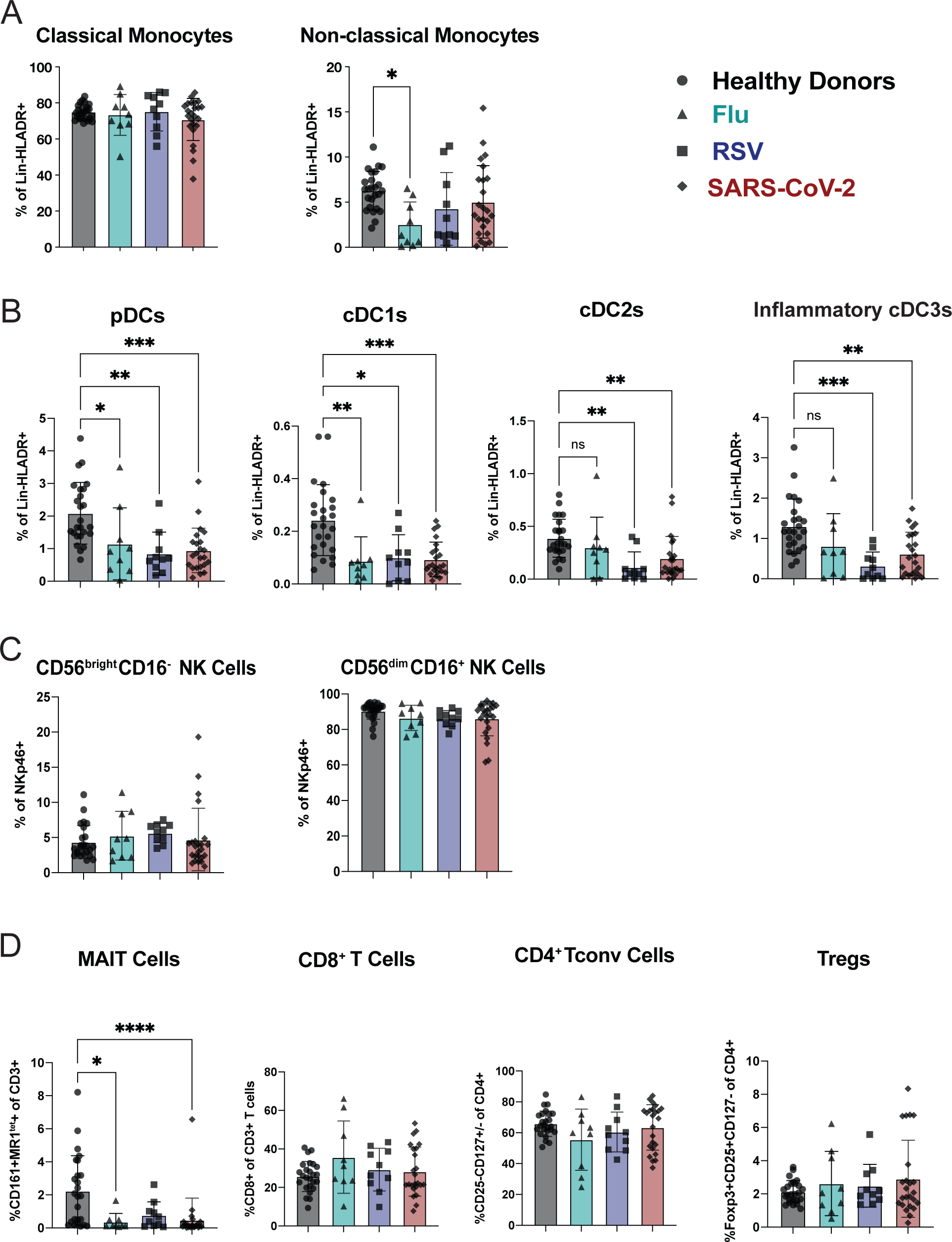
A decreased frequency of dendritic cell subsets is common across respiratory infections compared to healthy donors. Previously frozen PBMCs isolated from each group of the cohort were stained using 4 high parameter flow cytometry panels. Manual gating was used to estimate the frequencies of **A)** the monocyte family (Supplemental Figure 1A), **B)** the DC family (Supplemental figure 1A), the NK cell family (Supplemental Figure 1B), and, **D)** the T cell family (Supplemental Figure 1C), for each group of the cohort. All data include at least 9 patients per group (Table 1) and are represented as mean ± SD. Depending on the distribution of our data, statistical analyses were performed using either one-way ANOVA or Kruskal Wallis test. * P < 0.05; ** P < 0.01; *** P < 0.001, **** P <0.0001.

Finally, we observed a reduction in peripheral mucosal-associated invariant T (MAIT) cells across all respiratory infections (**Figure 2D** and **Supplemental Figure 1C**), similar to what has been previously documented in patients with severe COVID-19 compared to healthy donors^39^. We did not observe significant reductions compared to healthy donors in the T cell compartment of patients with any respiratory infection when we examined individual CD4 and CD8 T cell subsets (**Figure 2D**). Furthermore, there was no statistically significant difference in the frequency of CD25^+^CD127^-^Foxp3^+^ Tregs in patients with any of the respiratory infections compared to healthy donors (**Figure 2D**). Thus, in terms of overall immune cell subset distribution, we found that the NK cell family and T cell family remained unchanged between infected patients and healthy donors. We also found a congruent reduction in circulating DC subsets for Flu, RSV, and SARS-CoV-2 patients as compared to healthy donors. Overall, our results indicate that these immune cell changes are a general feature of immune responses to respiratory virus infections rather than a unique signature of SARS-CoV-2 infection.

### Immune cell phenotypic changes are consistent with both a respiratory virus signature as well as a SARS-CoV-2 specific signature

After observing minimal changes in the frequency of various immune cell subsets between respiratory viral infections, we next wanted to more comprehensively assess the expression of various markers of activation, maturation, and migration among monocytes, DC, NK cells, CD8^+^ T cells, CD4^+^ conventional T cells (Tconv), and CD4^+^ regulatory T cells (Tregs). Some markers did not show any change either across infection or compared to healthy donors and are displayed unabridged in **Supplemental Figures 2-4**. Interestingly, in instances wherein we observed a significant difference for one of the infections compared to healthy donors, this difference was usually seen across multiple respiratory infections, consistent with a respiratory virus signature (**Figure 3A-D**). Specifically, we observed in the monocyte population an increase in CD40 and CD206 expression in multiple, but not all, respiratory infections. Non-classical monocytes had a significant increase in CD11b and CD206 across all respiratory infections compared to healthy donors (**Figure 3A**). We observed few significant changes in the DC subsets, one being in the cDC3 population; the fraction of CD86-expressing cells was significantly lower in patients with Flu and RSV, while CD206 was significantly higher in patients with SARS-CoV-2 (**Figure 3B**). The frequency of pDCs expressing CD32 or CD38 was significantly increased in patients with any respiratory infection as compared to healthy donors (**Figure 3B**). In the two NK cell subsets, we observed a strong NK cell activation signature in patients hospitalized with respiratory viral infections, characterized by an increased frequency of CD38, CD69, HLA-DR, Ki67, and Granzyme B (**Figure 3C**). While others have shown this activated phenotype in NK cells following SARS-CoV-2 infection^13, 14^, we demonstrate that this phenotype is also a feature of NK cells in patients with other severe respiratory infections compared to healthy donors (**Figure 3C**).

**Figure 3.**
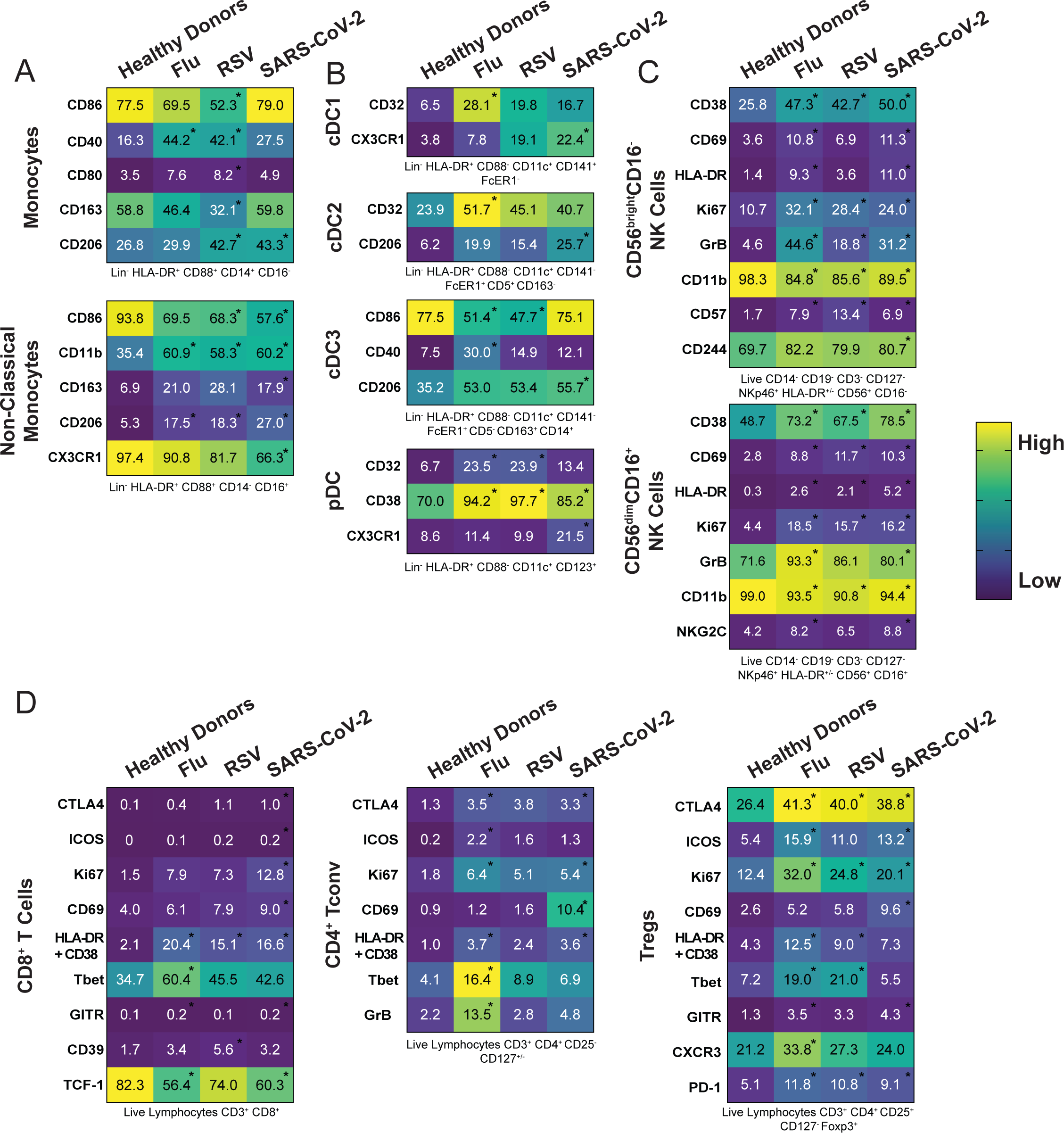
Immune cell phenotypic changes are consistent with a respiratory virus signature. (A-D) Heatmaps representing the expression pattern for all the indicated molecules within the main subsets of **A)** the monocyte family, **B)** the DC family, **C)** the NK cell family and, **D)** the T cell family for each group of the cohort. Gating strategy of the different subsets can be found under each heatmap (also see Supplemental Figure and numbers inside boxes represent the mean of frequency for each marker among the specific subsets. Depending on the distribution of our data, statistical analyses were performed using either one-way ANOVA or Kruskal Wallis test. Asterix inside the boxes is indicative of a significant difference compared to the healthy donors’ group and can include a p-value from 0.05 to 0.0001.

**Figure 4.**
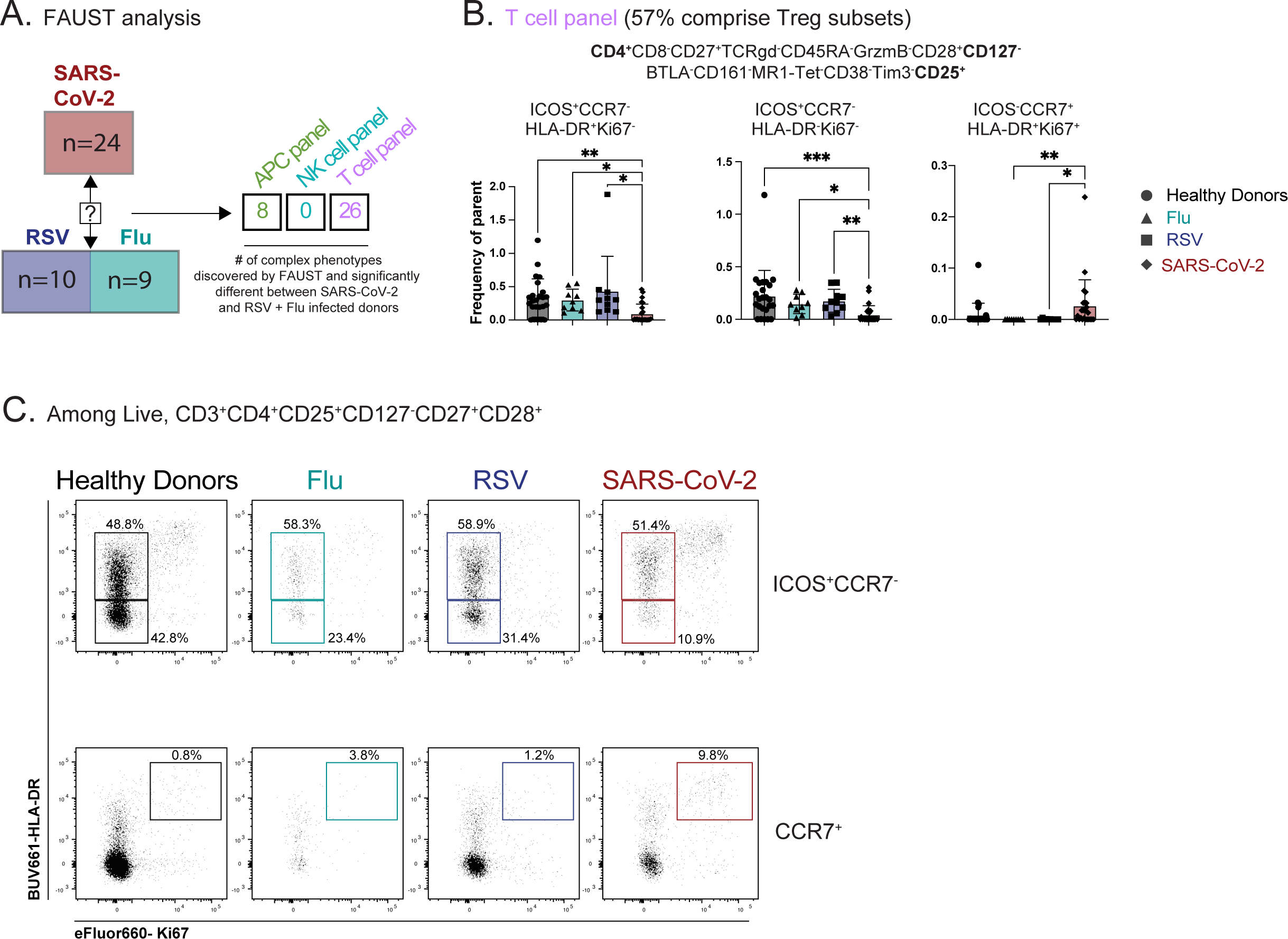
Unsupervised analysis reveals a SARS-CoV-2-specific signature including complex Treg phenotypes. **A)** FAUST analysis was used to discover complex phenotypes in the APC, NK and T cell panels. The multiple comparisons were adjusted using the Bonferroni correction and the numbers in the table are showing the number of identified phenotypes that are significantly different for SARS-CoV-2 infected patients compared to the Flu and RSV infected patients for each panel, with Bonferroni adjusted p-values under 0.05 considered significant. **B)** Example of three Treg phenotypes identified by FAUST within the T cell panel as shown in **Supplemental Table 2**. Bar graphs display the frequency of the phenotype for each group of the cohort among live, CD3^+^ cells. Data are represented as mean ± SD. Statistical analyses displayed were performed using Kruskal Wallis test. * P < 0.05; ** P < 0.01, *** P < 0.001. **C)** Representative flow plots showing the expression pattern of Ki67^+^ HLA-DR^+^cells among either CCR7^-^ICOS^+^ or CCR7^+^, live, CD3^+^CD4^+^CD25^+^CD127^-^CD27^+^CD28^+^ among the different groups of the cohort. Manual gating was performed using the T cell panel.

Finally, we found T cell subsets to have increased frequency of markers related to activation and effector function in individuals who were hospitalized for SARS-CoV-2 and other respiratory virus infections compared to healthy donors (**Figure 3D**). In particular, CD8 and CD4 Tconv cells positive for HLA-DR and CD38 were increased in patients hospitalized with Flu, RSV, and SARS-CoV-2 compared to healthy donors.

There was also an increase in the fraction of CD4 Tconv cells expressing CTLA-4 or Ki67 across all infections compared to healthy donors (**Figure 3D**). Finally, we also observed an increased frequency of Tregs expressing activation and suppression markers CTLA-4, ICOS, Ki67, HLA-DR/CD38, and PD-1 in patients with respiratory viruses compared to healthy donors (**Figure 3D**). It has been demonstrated during SARS-CoV-2 infection that NK and T cell subsets have increased activation and function compared to healthy donors^13, 14, 40^, and we hereby demonstrate that this phenomenon is not specific to SARS-CoV-2 infection, but rather spans multiple respiratory infections. We highlighted phenotypic marker alterations consistent between Flu, RSV, and SARS-CoV-2 infections compared to healthy donors, and we propose that these markers indicate a common circulating immune signature to respiratory virus infection.

### Unsupervised complex phenotype discovery analysis reveals a SARS-CoV-2 specific signature including complex Treg phenotypes

To specifically search for infection-specific changes in immune cell subsets in a unsupervised manner, we applied a recently developed non-parametric method for unbiased complex phenotype discovery called <u>F</u>ull <u>A</u>nnotation <u>U</u>sing <u>S</u>hape-constrained <u>T</u>rees (FAUST)^41^. Briefly, FAUST performs data-driven phenotype discovery and annotation on a per-sample basis, enabling the identification of statistically different complex immune phenotypes between the different groups of our cohort (**Supplemental Table 2**). We tested for differences in immune phenotypes between SARS-CoV-2 cohorts relative to Flu and RSV to determine if there were any complex immune cell phenotypes unique to SARS-CoV-2 infection using our high parameter flow panels (**Supplemental Table 1**).

Data from the APC panel revealed 8 distinct phenotypes with significant differences in cell frequency when comparing SARS-CoV-2 (all severity levels) to Flu and RSV (**Figure 4A** and **Supplemental Table 2**). There were no significant complex phenotypes discovered using the NK cell panel when patients with all SARS-CoV-2 severity levels were compared to patients with RSV or Flu (**Figure 4A** and **Supplemental Table 2**), in agreement with the NK cell analysis shown in **Figure 3** demonstrating that alterations in immune cell populations are largely consistent between respiratory infections. However, using the T cell panel in combination with FAUST analysis revealed 26 complex immune cell phenotypes that differed significantly in patients with SARS-CoV-2 infection with any level of disease severity compared to Flu and RSV (**Figure 4A**). Notably, the majority of these significantly different T cell phenotypes were CD4+CD25+CD127-, and thus comprising a Treg population (**Figure 4B** and **Supplemental Table 2**). Several subsets of CD4+CD25+CD127-Treg that were CD45RA-CCR7-(effector memory phenotype) were decreased in SARS-CoV-2 samples compared to the other respiratory infections and healthy donors, including a CD27+CD28+ICOS+HLA-DR+Ki67-and a CD27+CD28+ICOS+HLA-DR-Ki67-subset (**Figure 4B**). In contrast, a subset of CD4+CD25^+^CD127^-^ Treg that is CD45RA^-^CCR7^+^ (central memory phenotype) that co-expresses CD27, CD28, Ki67, and HLA-DR was significantly increased in the circulation of patients with SARS-CoV-2 and critical disease compared to Flu or RSV (**Figure 4B**). We confirmed these populations by manual gating of our flow cytometry data (**Figure 4C**). While the functional relevance of these cells is unclear, it is noteworthy that these complex Treg phenotypes distinguish SARS-CoV-2 infection compared to other respiratory infections, Flu and RSV. In summary, while much of the immune landscape is shared across these respiratory infections, our unsupervised analysis approach reveals unique complex phenotypes in the circulating Treg population that distinguishes patients infected with SARS-CoV-2 compared to Flu or RSV.

### Pro-inflammatory cytokines and chemokines are increased during respiratory infections compared to healthy donors

We next sought to determine if measuring cytokine and chemokine concentrations in the serum would provide additional insight to explain the overlap in immune phenotypes as well as the differences in Treg phenotypes. We tested serum samples from a subset of the cohort described in **Table 1** to quantify 71 different cytokines and chemokines (**Supplemental Table 3**). This analysis revealed a significant increase in serum IL-6 levels compared to healthy controls in both SARS-CoV-2 and RSV patients (**Figure 5A**). Others have shown an increase in IL-6 to be consistent with SARS-CoV-2 infection^7, 42, 43^, and here we demonstrate that IL-6 is significantly elevated in the serum of RSV patients as well. As IL-6 is an important pro-inflammatory cytokine during mucosal infections^44^, we wanted to examine whether other pro-inflammatory cytokines were increased during respiratory viral infections compared to SARS-CoV-2. We observed a significant increase in IL-8, IL-15, and IL-10 during both SARS-CoV-2 infection and RSV infection (**Figure 5A**). We did not see a large increase in pro-inflammatory cytokine levels in patients infected with Flu. However, this could be due to the reduced number of serum samples available in our cohort from Flu patients (N=3). Inflammatory mediators such as interferons (IFNs), IL-1α, and IL-1β have also been reported to be increased in patients with COVID-19^2^, although some reports have demonstrated very low levels of type I IFNs (IFNα and IFNβ)^6^. In our cohort of patients, we observed increased levels of IL-1β and IL-1α in patients with RSV compared to healthy donors, but no difference between healthy donors and SARS-CoV-2 or Flu patients. Furthermore, there was no difference in type I or type II interferons in patients infected with SARS-CoV-2 compared to healthy donors, although serum IFNα was significantly increased during RSV infection compared to either healthy donors or SARS-CoV-2 patients (**Figure 5A**). We also observed a significant elevation of IL-1RA during both SARS-CoV-2 and RSV infection (**Figure 5A**). Inflammatory chemokines were significantly increased during respiratory infection compared to healthy donors, with serum levels of CXCL9 and CXCL10 elevated during SARS-CoV-2 infection, though CXCL9 was also significantly increased in the context of RSV infection compared to healthy donors (**Figure 5B**). TRAIL has previously been correlated with viral load during SARS-CoV-2 infection^10^, but we found it to be significantly decreased in the serum of RSV and SARS-CoV-2 patients compared to healthy donors in our cohort (**Figure 5B**). Additionally, CCL17 and CCL22, chemokines known to be involved in the mobilization of immune cell to the lungs^45, 46^ and reported to be increased in SARS-CoV-2 infection^10, 47, 48^, were decreased or unchanged in the serum compared to healthy donors in our cohort. These findings are most likely due to the timing of the sample collection from symptom onset; studies have shown that trafficking of immune cells by these chemokines are most elevated as early as 1 week after infection^47^, and since all of our samples came from hospitalized patients, some were collected weeks after symptom onset (**Table 1**). Finally, due to variability in the quantities of these cytokines and chemokines detected in serum of SARS-CoV-2 patients, we next assessed whether levels of these cytokines and chemokines differed by severity of COVID-19, as defined by oxygen supplementation requirements (COVID-19 moderate, severe, or critical).

**Figure 5.**
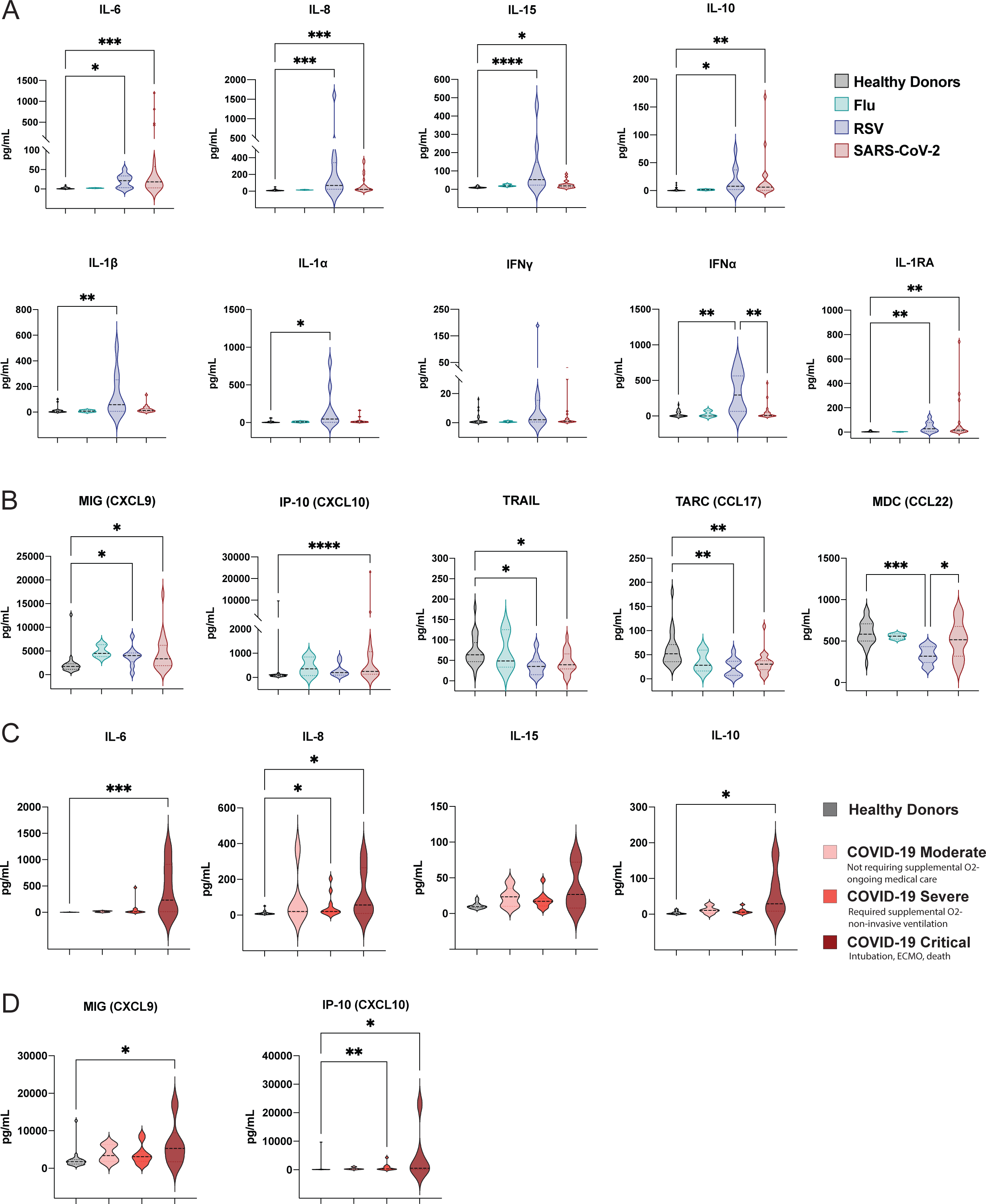
Pro-inflammatory cytokines and chemokines are increased during respiratory infections compared to healthy donors. **(A-B)** Violin plots showing the **A)** cytokine and **B)** chemokine concentrations in the serum for healthy donors (n=25), patients infected by Flu (n=3), RSV (n=10) and SARS-CoV-2 (n=22) (**C-D**) SARS-CoV-2 infected patients were grouped based on the severity of the disease (see Table 1) and Violin plots are showing the **C)** cytokine and **D)** chemokine concentrations in the serum for healthy donors as well as moderate COVID-19 (n=5), severe COVID-19 (n=11) and critical COVID-19 (n=6). All data are represented as mean ± SD. Depending on the distribution of our data, statistical analyses were performed using either one-way ANOVA or Kruskal Wallis test. * P < 0.05; ** P < 0.01; *** P < 0.001, **** P <0.0001.

Serum levels of IL-6, IL-8 and IL-10 were all significantly increased in patients with critical COVID-19 as compared to healthy donors, thereby suggesting that these cytokines are a feature of critical disease (**Figure 5C**). Additionally, the chemokines CXCL9 and CXCL10 were significantly elevated in patients with increased COVID-19 severity (**Figure 5D**). Overall, we demonstrated that several cytokines and chemokines previously associated with SARS-CoV-2 infection were also elevated in the serum of patients hospitalized with other respiratory infections, and so may not be a unique feature of COVID-19.

### Markers of cellular activation among NK and T cells are increased after COVID-19 to varying degrees

Based on the increase in pro-inflammatory cytokines and chemokines with increasing COVID-19 disease severity, we wanted to determine if effector immune cell subsets were also altered with COVID-19 severity in our cohort. In the CD56^bright^CD16^-^ population of NK cells, characterized as being cytokine producers with proliferative potential^49^, we observed an increased expression of both CD38 and CD69 for both patients with severe and critical COVID-19 compared to healthy donors (**Figure 6A**). We also observed an increased expression of Ki67 among CD56^bright^CD16^-^ NK cells of patients with severe COVID-19 compared to healthy donors.

**Figure 6.**
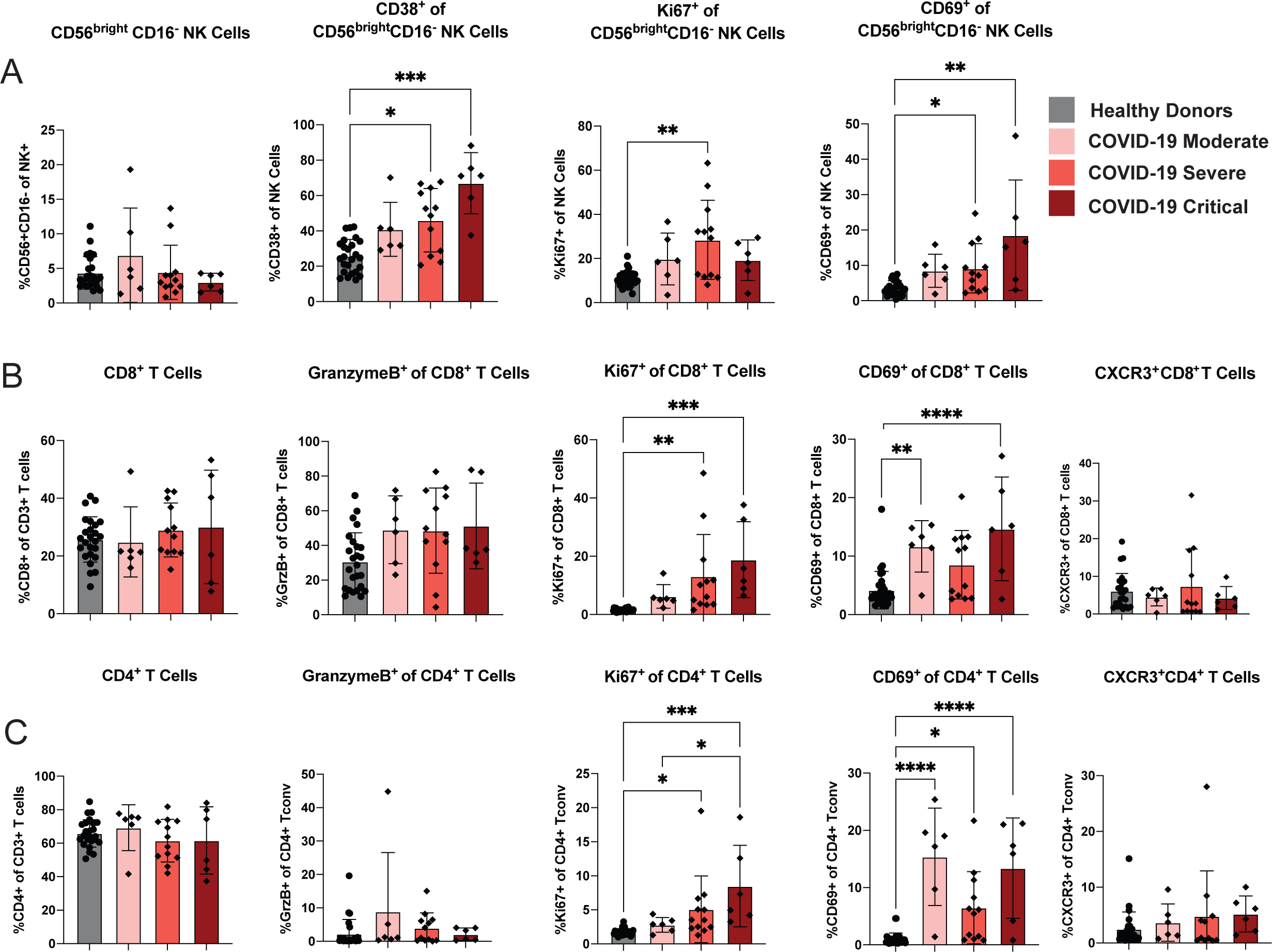
Markers of cellular activation among NK and T cells are increased after COVID-19 to varying degrees. SARS-CoV-2 infected patients were grouped based on the severity of the disease and markers of cellular activation were analyzed among **A)** CD56^bright^ CD16^-^ NK cells **B)** CD8^+^ T cells and **C)** CD4^+^T cells, for healthy donors as well as moderate COVID-19 (n=6), severe COVID-19 (n=12) and critical COVID-19 (n=6). All data are represented as mean ± SD. Depending on the distribution of our data, statistical analyses were performed using either one-way ANOVA or Kruskal Wallis test. * P < 0.05; ** P < 0.01; *** P < 0.001, **** P <0.0001.

Upon examination of T cell activation status, we observed that the frequency of CD8^+^ T cells was not altered based on COVID-19 severity, nor was the frequency of CD8^+^ T cells expressing the cytotoxic molecule granzyme B (**Figure 6B**). However, the frequency of CD8^+^ T cells expressing Ki67 was significantly elevated for both patients with severe and critical COVID-19 (**Figure 6B**). We also found an increased CD69 expression among circulating CD8^+^ T cells in both patients with moderate and critical COVID-19 (**Figure 6B**). Finally, CXCL9 and CXCL10 are inflammatory chemokines induced by IFNγ that share the chemokine receptor CXCR3^50^. Since these chemokines were increased in patients with severe COVID-19 (**Figure 5D**), we next wanted to determine if CXCR3 expression was altered on T cells, thereby potentially accounting for the increased fraction of activated cells present in the circulation of patients with critical COVID-19. However, the expression of CXCR3 was not significantly increased by CD8^+^ T cells from patients with any degree of COVID-19 severity (**Figure 6B**). A similar expression pattern of activation markers was observed in CD4^+^ Tconv cells; there was no change in the frequency of CD4^+^ T cells based on COVID-19 severity, and there was limited expression of granzyme B within the CD4^+^ T cell subset that did not vary by disease severity. However, the frequency of CD4^+^ T cells expressing either Ki67 or CD69 was increased in patients with COVID-19, with Ki67 increasing with disease severity (**Figure 6C**). We did not see any increase in the fraction of CD4^+^ T cells that expressed CXCR3 (**Figure 6C**), suggesting that these activated T cells would not have the potential to migrate to the lung using the mucosal tissue homing molecule CXCR3, though they may utilize other chemokine receptors to enter this critical site of virus replication. We sought to confirm that any increase in markers of cellular activation were in fact due to COVID-19 severity and not related to days post-symptom onset. When we examined these markers of cellular activation on NK cells and T cells by days post symptom onset to sample collection, we found no significant difference in cellular activation relating the days post-symptom onset in our SARS-CoV-2 cohort (**Supplemental Figure 5**). This suggests that our findings are associated with COVID-19 severity rather than timing of sample collection.

### Regulatory T cells in patients with critical COVID-19 disease are increased in frequency and display a heightened activation signature

A hallmark of the immune response to SARS-CoV-2 infection in individuals with severe disease has been identified as a state of dysregulated and pro-inflammatory immunity^6– 8, 10, 13, 14, 28, 32, 40, 51, 52^. We and others have previously demonstrated that Tregs play a role in orchestrating the anti-viral immune response by potentiating the antigen-specific T cell response^53–58^. However, it is also evident that in the context of infections, including RSV and Flu, Tregs can assist in restraint of immunity to reduce immunopathogenesis associated with a robust immune response^54, 58, 59–64^. Because we identified significant changes in the frequency of complex Treg phenotypes in patients with SARS-CoV-2 compared to Flu and RSV infection (**Figure 4**), we sought to further examine Treg phenotype based on COVID-19 severity. Since we also observed increased proliferation of CD4^+^ Tconv cells with increased COVID-19 severity, we hypothesized that Tregs could be involved in restraining this exuberant anti-SARS-CoV-2 immune response. A previous study found no significant difference in the frequency of Tregs in the circulation by COVID-19 severity^7^, while two subsequent studies have identified a slight decrease in Treg frequency with increasing COVID-19 severity^11, 26^. However, when we measured Treg frequency in healthy donors compared to patients with COVID-19 disease, we saw a significant increase in the frequency of CD25^+^CD127^-^Foxp3^+^ Tregs in COVID-19 critical patients only (**Figure 7A**). We additionally measured the median fluorescent intensity of Foxp3 in the Tregs, as this has been shown to be an indicator of suppressive capabilities^65^. We detected an increase in the level of Foxp3 expression by Tregs in the COVID-19 critical patients compared to patients with moderate disease (**Figure 7A**). To test if circulating Tregs from COVID-19 patients showed a more suppressive phenotype, we measured Ki67, CTLA-4, GITR, and ICOS, all of which were significantly increased with COVID-19 disease severity (**Figure 7B**). TCF1, a transcription factor that has been shown to dampen Foxp3 activity^66, 67^, appeared to be decreased with COVID-19 severity, albeit non-significantly, consistent with the notion of more functional Tregs in COVID-19 critical patients (**Figure 7B**). Finally, we wanted to determine if Tregs were licensed to migrate to the lungs in COVID-19 patients, and so we examined CXCR3 expression. We observed a significant increase in the frequency of CXCR3 expressing Tregs with increasing COVID-19 severity (**Figure 7B**). Our data indicated that Tregs are highly activated in patients with critical COVID-19 and are potentially able to migrate toward a gradient of increasing CXCL9 and CXCL10 during SARS-CoV-2 infection.

**Figure 7.**
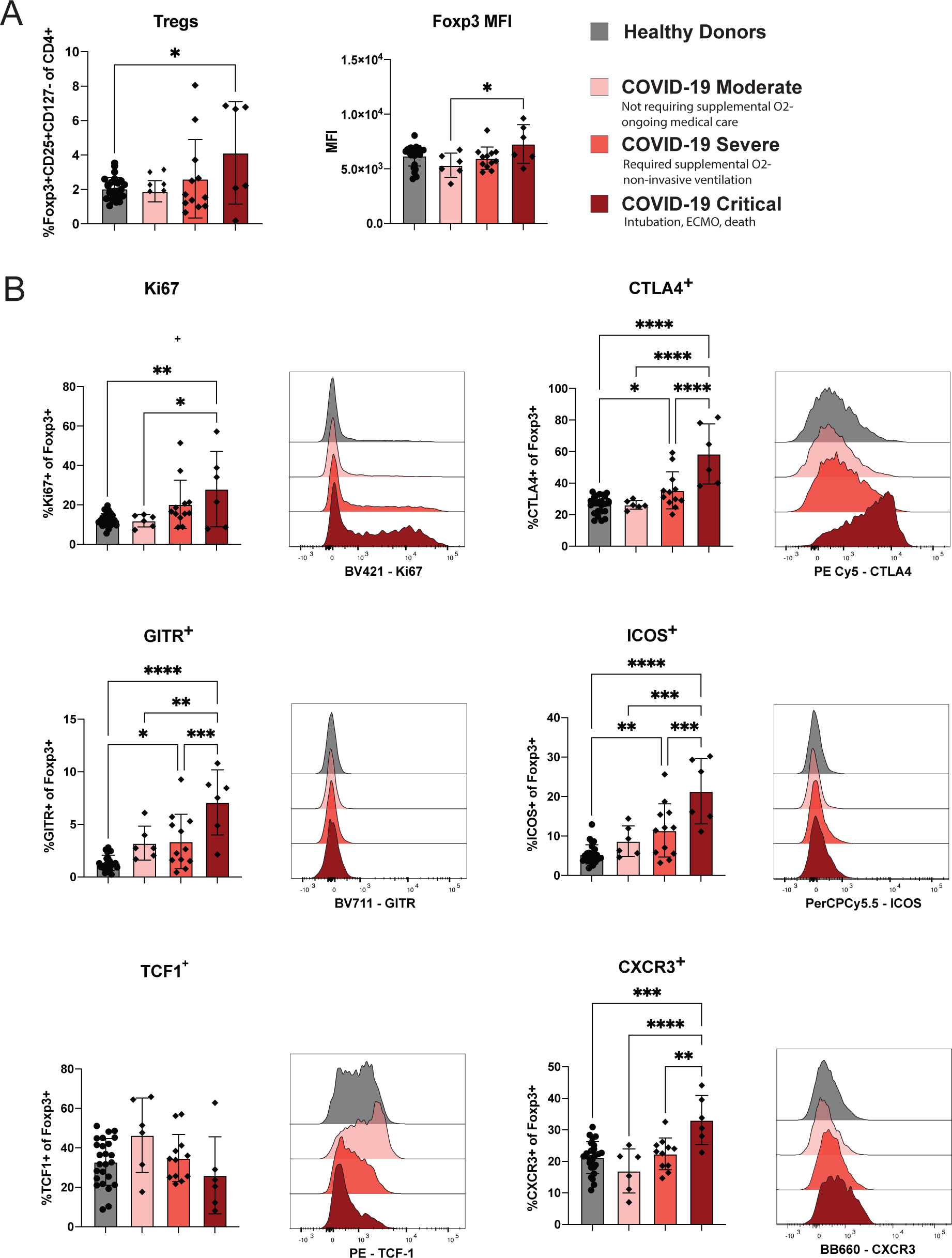
Regulatory T cells in patients with critical COVID-19 disease are increased in frequency and display a heightened activation signature. **A.** Bar graphs showing (*left*) the frequency of Treg and (right) the MFI of Foxp3 among parent for healthy donors and severity-based groups of COVID-19. **B.** Representative histograms and quantification of the expression of activation and suppressive markers within Tregs for healthy donors as well as moderate COVID-19 (n=6), severe COVID-19 (n=12) and critical COVID-19 (n=6). All data and are represented as mean ± SD. Depending on the distribution of our data, statistical analyses were performed using either one-way ANOVA or Kruskal Wallis test. * P < 0.05; ** P < 0.01; *** P < 0.001, **** P <0.0001.

## Discussion

More than a year into the COVID-19 pandemic, numerous studies of peripheral blood from individuals infected with SARS-CoV-2 have revealed that a hyper-inflammatory and dysregulated immunotype is characteristic of COVID-19 patients compared to healthy donors. In attempts to identify unique aspects of anti-SARS-CoV-2 immunity that could underlie disease presentation and severity, a comparison with other common respiratory virus infections is required. However, there have only been a limited number of studies comparing immune phenotypes generated after SARS-CoV-2 infection to other respiratory viral infections^6, 28, 68^. It was first demonstrated by Blanco-Melo *et al* that compared to other respiratory viral infections, including human parainfluenza virus 3 and RSV, SARS-CoV-2 elicits a blunted early type I and type III interferon response *in vitro* and in animal models^6^. Through an scRNAseq study of PBMCs from individuals with COVID-19 or severe influenza, another group demonstrated that cells from COVID-19 patients had a predominantly IL-1β and TNF inflammatory signature, whereas Flu patients had an increased interferon-stimulated gene (ISG) response^68^, thereby uncovering differential pro-inflammatory pathways elicited by distinct respiratory viral infections. Finally, a recent study comparing immune responses in patients with severe influenza or COVID-19 found that the latter exhibited similar lymphocyte counts but fewer monocytes and reduced HLA-DR expression on monocyte subsets as compared to Flu patients^28^. To extend these studies, we designed a study to comprehensively examine serum cytokines and chemokines as well as the immunotypes of myeloid cells, NK cells, T cells, and Tregs in the peripheral blood of patients hospitalized with Flu or RSV compared to SARS-CoV-2 or healthy controls. Importantly, we used high-parameter flow cytometry coupled with both unbiased computational analysis approaches as well as traditional manual gating to perform a comprehensive examination of many immune cell subsets as well as complex phenotypes. We reasoned that comparison of immune phenotypes between patients hospitalized with COVID-19 versus other severe respiratory virus infections could potentially reveal common immunotherapeutic strategies that can thus be leveraged in the battle against SARS-CoV-2 and future pandemic viruses. For example, knowledge of immune-targeting therapeutic strategies to treat SARS-CoV-2 could potentially be applied to the next pandemic respiratory virus, which may be Flu or another CoV.

Analysis of our cohort demonstrates that most of the previously identified alterations in peripheral immune populations during SARS-CoV-2 infection are not a distinguishing features of the anti-SARS-CoV-2 immune response, but rather indicate a more common immune landscape associated with respiratory viruses in general. We have identified a general respiratory virus-induced immune signature across three different respiratory viral infections compared to healthy donors (**Figure 3A-D**).

However, applying an unsupervised phenotype discovery analysis (FAUST) revealed previously undescribed alterations of Tregs with various complex phenotypes in SARS-CoV-2 patients compared to those with Flu or RSV (**Figure 4**). More specifically, we detected a reduced frequency of effector memory phenotype (CD45RA-CCR7-) Tregs co-expressing various markers of activation, including ICOS, CD27, CD28, and HLA-DR present in the blood of patients with SARS-CoV-2 compared to Flu or RSV (**Figure 4B- C**). This could reflect a reduction in activated Tregs able to migrate to the peripheral tissues including the lung, whereby they could participate in restraining immunopathology and limiting ARDS. In contrast, we identified a unique population of blood Tregs with a central memory phenotype (CD45RA^-^CCR7^+^) co-expressing CD27, CD28, Ki67 and HLA-DR that was present at a significantly increased frequency in SARS-CoV-2 patients compared to those with Flu or RSV (**Figure 4B-C**). We speculate that these Tregs represent a circulating population of activated suppressive cells that may participate in restraining the inflammatory response in the context of COVID-19. However, whether or not this is of benefit to the host in the context of disease is an open question. Thus, additional studies are required to determine if Treg-modulating therapies could be of benefit in limiting COVID-19 severity. We recently demonstrated that in a mouse model of SARS-CoV, an elevated steady-state, pre-infection frequency of Tregs correlates with protection from high viral loads and disease upon infection^69^, thereby contributing to the notion that Tregs could play a protective role in limiting disease. However, examination of prospectively collected pre-COVID-19 pandemic samples from humans that went on to become infected would be required to establish whether or not Treg abundance is predictive of viral loads or disease severity upon SARS-CoV-2 infection. Recent evidence suggests that the airways of patients with severe COVID-19 have reduced Treg frequencies compared to healthy airways^27^, leading us to speculate that there may be a defective trafficking of Tregs from the circulation into the respiratory tract in the context of COVID-19, thus contributing to lung immunopathogenesis. We hypothesize that this may be due to the increased levels of CXCL10 and CXCL9 found in peripheral blood (**Figures 5B and 5D**) that may retain CXCR3^+^ Treg in the periphery and prevent them from entering the airways via a chemokine gradient. In addition, the reduced frequency of effector memory phenotype Treg present in the blood of patients infected with SARS-CoV-2 compared to Flu and RSV may indicate that Treg able to migrate to tissue sites may be diminished in the context of SARS-CoV-2. Thus, while additional studies of the mucosal immune response to SARS-CoV-2 are warranted, we speculate that immunotherapies designed to attract Treg out of the circulation and into the respiratory tract could be beneficial in limiting disease. Of note, there are several ongoing clinical trials that target various chemokine receptors, including CCR2 and CCR5, in an effort to minimize immune-mediated lung tissue damage (NCT04435522 and NCT04500418).

Our study has some limitations, first of which is our exclusive focus on peripheral blood immune responses rather than tissue-specific responses. In addition, our cohort includes patient sample collection from variable times post-symptom onset, from zero to 47 days. This variability could clearly impact the types of immune phenotypes detected, as could variability in viral loads and durability of viral shedding, for which we are lacking data from the majority of patients due to scarcity of testing in the early days of the pandemic. Finally, while we were powered to uncover unique aspects of the circulating Treg phenotypes of patients with SARS-CoV-2 compared to Flu or RSV, our relatively small N (**Table 1**) may have precluded identification of other distinguishing immune phenotypes.

In sum, our study based on high dimensional flow cytometry data combined with several analysis methods reveals a largely similar immune landscape of patients hospitalized with respiratory virus infections, including SARS-CoV-2. This is further supported by our analysis of 71 soluble cytokines and chemokines in the blood of patients with SARS-CoV-2, Flu, or RSV. The recent identification of novel SARS-CoV-2 variants that may increase transmission and alter vaccine efficacy^70^ underscores the need for continued development of treatment strategies specifically for severe COVID-19 disease course. Thus, we speculate that the overlapping immune landscapes in SARS-CoV-2, Flu, and RSV infections could be leveraged to identify and hasten common treatment strategies that could be leveraged for the response to the next pandemic respiratory virus. Surprisingly, we identified that SARS-CoV-2 patients with the most critical disease presented with unique alterations in the Treg compartment, including an increase in a population of CD45RA^-^CCR7^+^Ki67^+^HLA-DR^+^ Tregs within the circulation compared to patients with Flu or RSV, and an increase in CXCR3^+^ Tregs in the blood of patients with COVID-19, thus leading us to predict that Treg-targeting therapies could be useful in limiting disease associated with SARS-CoV-2. Additional studies of Tregs present in the respiratory tract of COVID-19 patients, as well as investigations into immune-therapeutic approaches to target multiple respiratory virus infections will be useful in identifying additional therapeutic avenues that can curtail viral infection-mediated severe lung disease, including disease induced by future pandemic viruses.

## Methods

### Sample Collection

#### Study Population

Study samples were collected as part of the prospective longitudinal cohort study HAARVI (Hospitalized or Ambulatory Adults with Respiratory Viral Infections) in Seattle, Washington. Individuals 18 years or older were eligible for inclusion and were recruited from two groups: inpatients with laboratory confirmed respiratory viral infection, and healthy controls. Inpatients were hospitalized at Harborview Medical Center, University of Washington Medical Center, or Northwest Hospital and identified through a laboratory alert system. A cohort of healthy individuals were enrolled in this study and were recruited through email and flyer advertising. They were considered eligible if they had no history of laboratory confirmed SARS-CoV-2 infection and had not presented with ILI (influenza-like illness) in the last 30 days.

Participants or their legally authorized representatives completed informed consent. Sociodemographic and clinical data were collected from electronic chart review and from participants via a data collection questionnaire (Project REDCap) ^71^ at the time of enrollment. The questionnaire collected data on the nature and duration of symptoms, medical comorbidities, and care-seeking behavior. Based on these data, individuals were classified by disease severity utilizing an eight-point ordinal clinical assessment scale ^72^. For our study, a clinical assessment score of 1 (death) or 2 (intubation, ECMO) was categorized as critical COVID-19. A score of 3 (non-invasive ventilation or high flow O2 device) or 4 (required supplemental O2) was categorized as severe COVID-19. Finally, a score or 5 (not-requiring supplemental O2) or 6 (no longer requires ongoing medical care) was categorized as Moderate COVID-19. There were no participants with a clinical assessment score of 7 or 8 because our cohort solely consisted of hospitalized patients. All SARS-CoV-2 patient samples were collected after March 1, 2020. All Flu and RSV patient samples were collected between 2017 and 2019 during Flu season.

#### Ethics

The studies were approved by the University of Washington Human Subjects Institutional Review Board, IRB numbers STUDY00000959 and STUDY00002929.

### Sample Processing

Participant samples obtained before March 1, 2020 were collected in Mononuclear Cell Processing (CPT, BD) and serum tubes and immediately transferred to the University of Washington. Whole blood in serum tubes was allowed to clot by incubating for at least 1 hour at room temperature then centrifuged at 700xg for 15 minutes, aliquoted, and stored at -20°C. CPT tubes were incubated for 2 hours at room temperature before centrifuging at 2000xg for 40 minutes. Purified PBMCs were transferred to a 15mL conical tube, washed twice with PBS, resuspended in Recovery Freezing Medium (Thermo Fisher Scientific, Waltham, MA), and stored in liquid nitrogen until use.

Participant samples obtained after March 1, 2020 were collected in acid citrate dextrose and serum-separating tubes (SST, BD) and immediately transferred to the University of Washington. Whole blood in SST tubes was allowed to clot by incubating for at least 1 hour at room temperature then centrifuged at 700xg for 10 minutes, aliquoted, and stored at -20°C. Peripheral blood mononuclear cells (PBMC) were isolated by density-gradient centrifugation using Histopaque (Sigma-Aldrich, St. Louis, MO). After washing, purified PBMC were resuspended in 90% heat-inactivated fetal bovine serum (FBS) (Sigma-Aldrich, St. Louis, MO) with 10% dimethyl sulfoxide (DMSO) (Sigma-Aldrich, St. Louis, MO) cryopreservation media and stored in liquid nitrogen until use. All samples were frozen within six hours of collection time.

### Flow Cytometry

For flow cytometric analysis, good practices were followed as outlined in the guidelines for use of flow cytometry ^73^. Directly following thawing, cells were incubated with Fc-blocking reagent (BioLegend Trustain FcX, #422302) and fixable UV Blue Live/Dead reagent (ThermoFisher, #L34961) in PBS (Gibco, #14190250) for 15 minutes at room temperature. After this, cells were incubated for 20 minutes at room temperature with 50 μl total volume of antibody master mix freshly prepared in Brilliant staining buffer (BD Bioscience, #563794), followed by two washes. All antibodies were titrated and used at optimal dilution, and staining procedures were performed in 96-well round-bottom plates. A detailed list of the main panels used, including Fluorochromes and final dilutions of all antibodies is provided in **Supplemental Table 1.**

The stained cells were fixed with 4% PFA (Cytofix/Cytoperm, BD Biosciences) for 20 minutes at room temperature, washed, resuspended in FACS buffer and stored at 4°C in the dark until acquisition. For panels with intranuclear staining, the cells were fixed with intranuclear transcription factor staining kit (eBioscience Foxp3/Transcription Factor Staining Buffer Set, Thermo Fisher #00-5532-00) following manufacturers’ protocols.

Single-stained controls were prepared with every experiment using antibody capture beads diluted in FACS buffer (BD Biosciences anti-mouse, #552843, anti-rat, #552844, and Miltenyi anti-REA, #130-1040693). Beads (ArC^TM^ Amine Reactive Compensation Bead Kit, Themo Fisher #A10346) or cells were used for Live/Dead single-stained control, and treated exactly the same as the samples (including fixation procedures). All samples were acquired using a FACSymphony A5 (BD Biosciences), equipped with 30 detectors and 355nm (65mW), 405nm (200mW), 488nm (200mW), 532nm (200mW) and 628nm (200mW) lasers and FACSDiva acquisition software (BD Biosciences). Detector voltages were optimized using a modified voltage titration approach ^74^ and standardized from day to day using MFI target values and 6-peak Ultra Rainbow Beads (Spherotec, # URCP-38-2K) ^75^. After acquisition, data was exported in FCS 3.1 format and analyzed using FlowJo (version 10.7.x, BD Biosciences). Doublets were excluded by FSC-A vs FSC-H gating.

Importantly, as the samples were stained and acquired in two different batches, each experiment was conducted along with a technical control: a cryopreserved vial of PBMC collected via leukapheresis from one single healthy donor. This method is valuable in order to ensure that the variability of expression of the different markers is neither due to variability on the instrument side nor staining-related and allows to compare data from biological samples stained on different days. For samples acquired on different experiment days, files were exported as compensated data and analyzed combined together in a new workspace. Gates were kept the same across all samples except where changes in the density distribution clearly indicated the need for adjustment.

### Cytokine and Chemokine Measurements

Patient serum aliquots were stored at −80 °C. Serum samples were shipped to Eve Technologies (Calgary, Alberta, Canada) on dry ice, and levels of cytokines and chemokines were measured using the Human Cytokine Array/Chemokine Array 71-403 Plex Panel (HD71). All samples were measured upon the first thaw.

### Statistical Analysis

After testing the normal distribution of our data using the D’Agostino & Person test, statistical analyses were performed using either an ordinary one-way ANOVA (parametric test) or Kruskal Wallis test (nonparametric test) using the GraphPad Software. Data are expressed as mean ± SD. Significant P values were annotated as follows. * P < 0.05; ** P < 0.01; *** P < 0.001; **** P < 0.0001.

## FAUST Analysis

FAUST was used to discover and annotate phenotypes in the 4 tested panels. Manual gating was first used in order to define on which cell type FAUST should be run. FAUST was applied to live cells for the APC panel; live, CD3^+^ cells for the T cell panel; live, CD3^+^ CD4^+^ CD25^+^ CD127^-^ for the Treg panel; and live, CD14^-^ CD19^-^ CD3^-^ CD127^-^ for the NK panel. After incorporating expert information about the panel design, FAUST selected markers to be used for annotation and discovery of phenotypes. The markers identified for the different panels were the following:

**APC panel**: CD1c, CD5, CD11b, CD11c, CD14, CD16, CD32, CD38, CD46. CD85k, CD86, CD88, CD123, CD141, CD163, CD301, CX3CR1, FcER1, PD-L1 and Sirpa

**T cell panel:** BTLA, CCR7, CD4, CD8, CD25, CD27, CD28, CD38, CD45RA, CD127, CD161, Granzyme B, HLA-DR, ICOS, Ki67, MR1-tet, TCRgd, Tim3

**Treg cell panel:** CCR5, CCR7, CD39, CD101, CTLA-4, CXCR3, GITR, KI67, Tbet, TCF-1

**NK cell panel**: CD2, CD16, CD38, CD56, CD57, CD69, Granzyme B, NKG2A, NKG2C, Ki67.

Then, within a given panel, a binomial generalized linear mixed-effects model (GLMM) with a subject-level random effect was used to test for association between counts of the discovered phenotypes and COVID-19 patients relative to Flu and RSV patients.

The severity of the COVID-19 disease was also tested in comparison to the other groups of the cohort (i.e., when the moderate COVID-19 patients were tested in comparison to Flu and RSV patients, the severe and critical COVID-19 patients were removed from the analysis). The entire collection of tested hypotheses was then adjusted using the Bonferroni adjustment. Within a given panel, discovered FAUST phenotypes with a Bonferroni adjusted p-values under 0.05 were selected.

### FlowSOM, UMAP, and Heatmap Generation

Pipelines outlined in the Spectre R package were used to generate UMAP and FlowSOM clusters ^29, 30, 76^. In FlowJo from the APC Panel, cells were gated by time (Time, FSC-A), cell size (SSC-A, FSC-A), singlets (FSC-H, FSC-A), and live (SSC-A, Dead). Each sample was then downsampled to 20,000 events if able and then exported as CSV files for the channel values of all parameters. Channel values in R were transformed by ArcSinH using a cofactor of 1000. Batches were normalized based on a reference sample run on both days using CytoNorm^76^. For clustering with FlowSOM and dimensionality reduction with UMAP, only lineages markers CD3, CD19, HLADR, CD56, CD14, CD88, CD16, CD11c, CD123, CD141, FcER1, CD5, and CD163 were used.

FlowSOM was initiated on default parameters where the number of clusters was automatically generated and produced 5. Clusters were then re-annotated to four labeling NK cells, T cells, B cells, and Lin-HLA-DR +/- cells. A UMAP was generated with default parameters on 40,000 events from downsampling 10,000 events per study group (healthy, RSV, FLU, and Sars-CoV2) so as to be equally represented. A heatmap was generated representing the median fluorescent intensity of the lineage markers normalized between 0 and 1 for the annotated clusters.

## Data Availability

We provide much of the raw data in the form of Supplementary Tables. Additional data can be provided upon request.

## Acknowledgements

We thank the members of the Lund and Prlic labs for helpful discussions, the HAARVI study team, and the patients and healthy donors for providing samples.

---

**Supplemental Figure 1.**
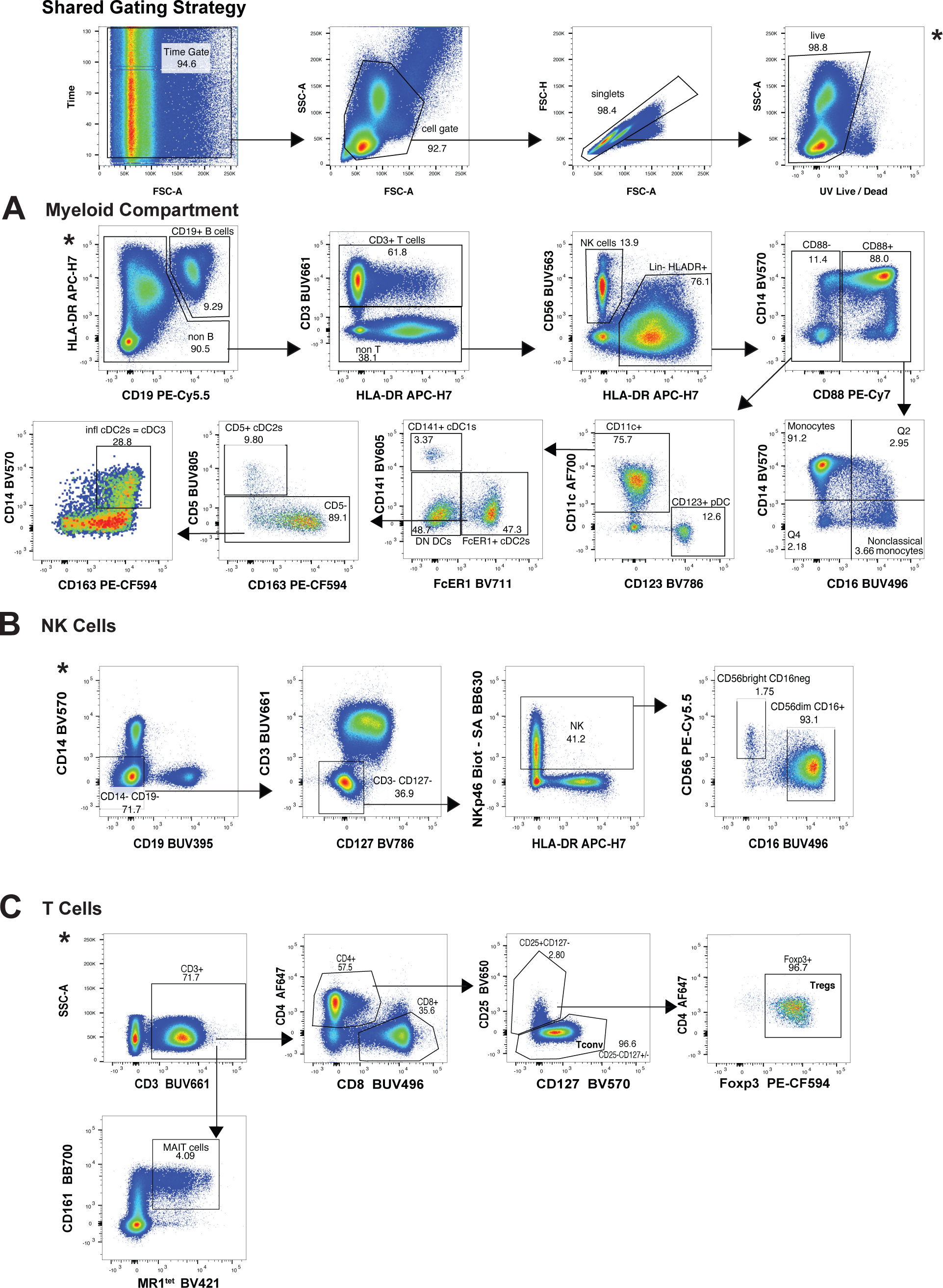
Gating strategies. Gating scheme used to identify **A)** the subsets of myeloid cells using the APC panel; **B)** the 2 NK cell subsets using the NK panel; and **C)** the T cell subsets using the Treg panel.

**Supplemental Figure 2.**
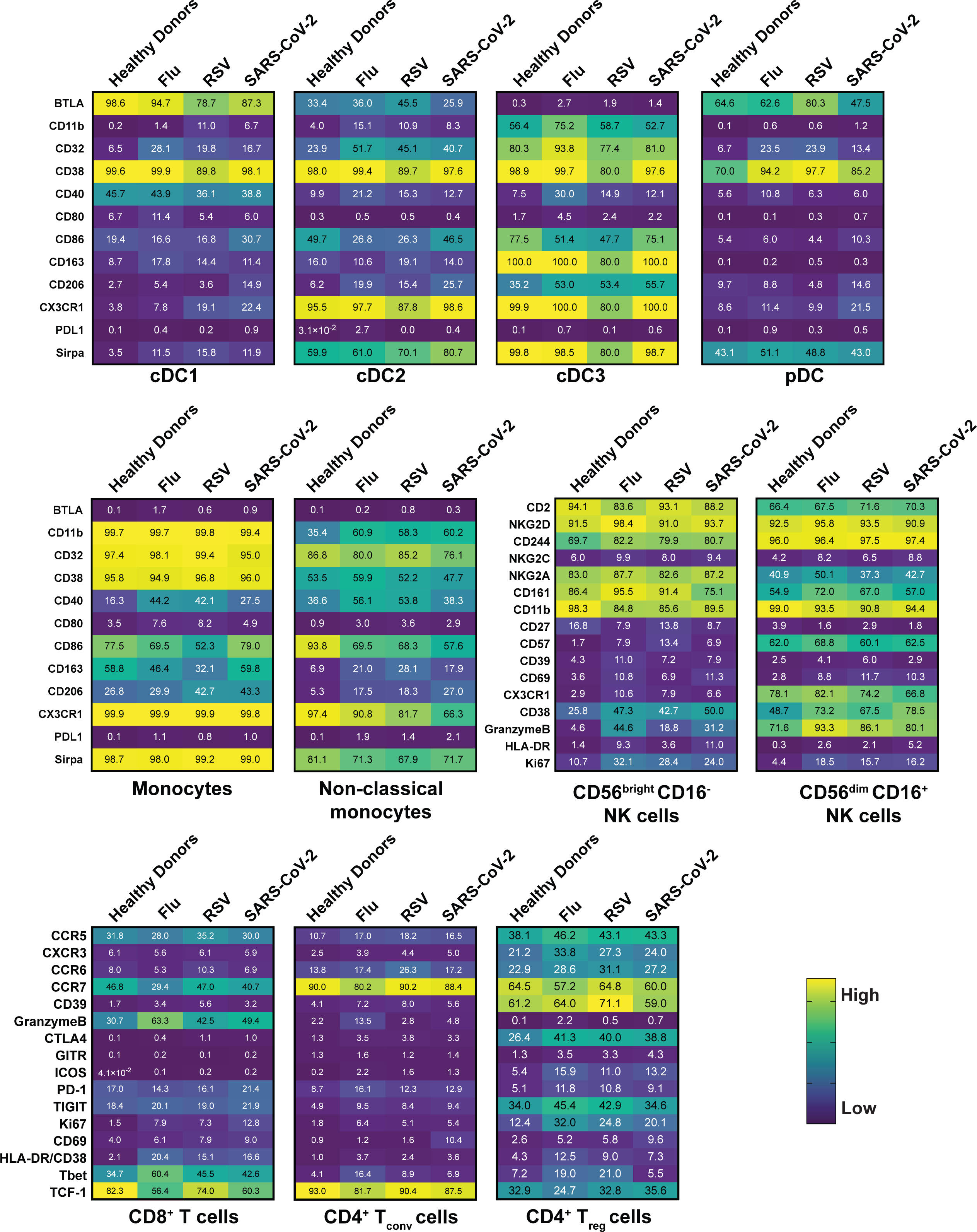
Similar and specific patterns of phenotypical changes among immune cells. Heatmaps representing the frequency for all the indicated molecules within the main subsets of **A)** the monocyte family, **B)** the DC family, **C)** the NK cell family and, **D)** the T cell family. Gating strategy can be found in Supplemental Figure 1 and numbers inside boxes represent the mean of frequency for each marker among the specific subsets.

**Supplemental Figure 3.**
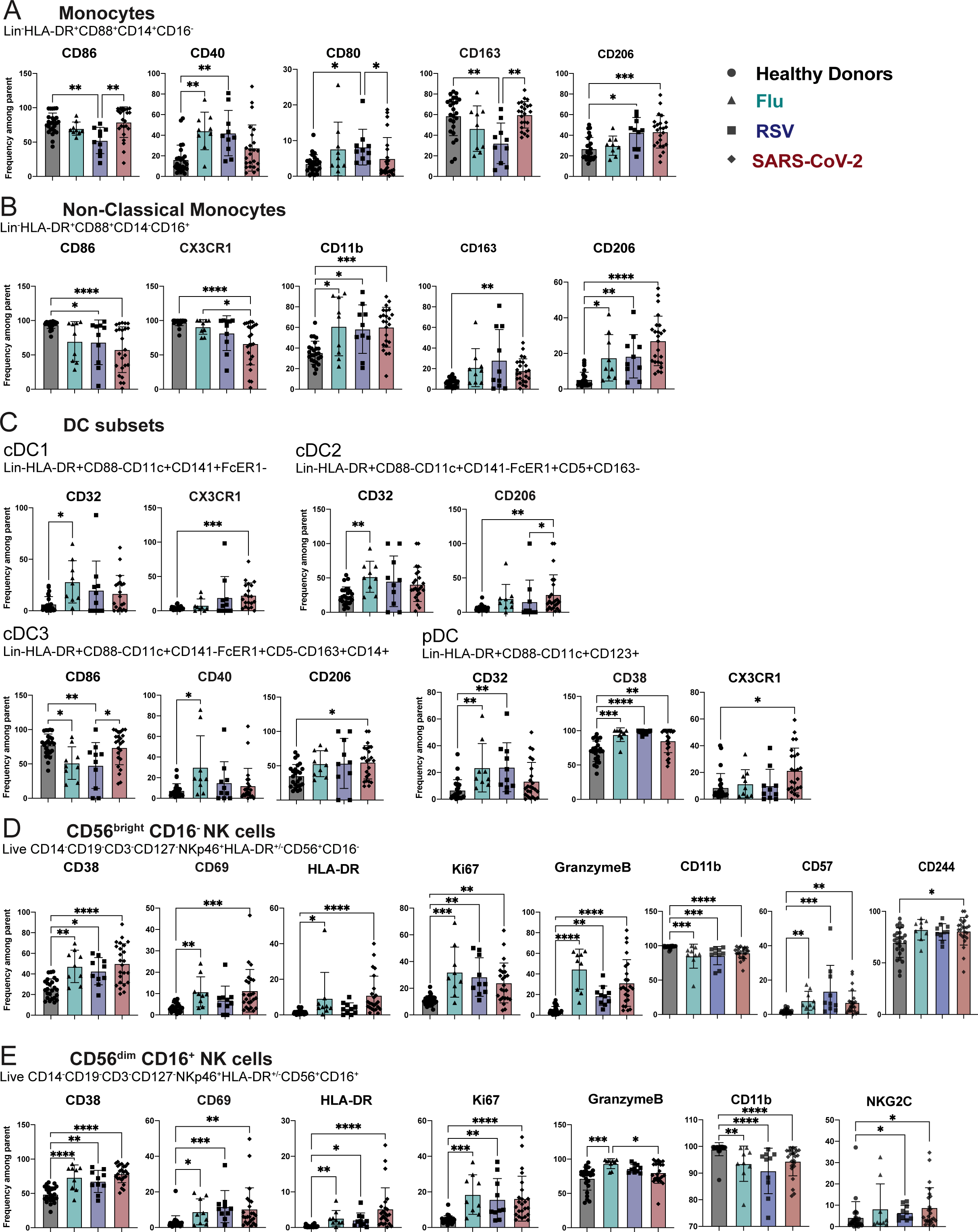
Phenotypical changes in the Myeloid and NK cell populations across respiratory infections. Bar graphs showing the mean frequency of indicated makers within the **A.** the monocyte, **B.** the nonclassical monocyte, **C.** the DC subsets, **D**. the CD56^bright^ CD16^neg^ NK cells, **E.** CD56^dim^ CD16^+^ NK cells. Gating strategy can be found in Supplemental Figure 1. All data include at least 9 patients per group (Table 1) and are represented as mean ± SD. Depending on the distribution of our data, statistical analyses were performed using either one-way ANOVA or Kruskal Wallis test. * P < 0.05; ** P < 0.01; *** P < 0.001, **** P <0.0001.

**Supplemental Figure 4.**
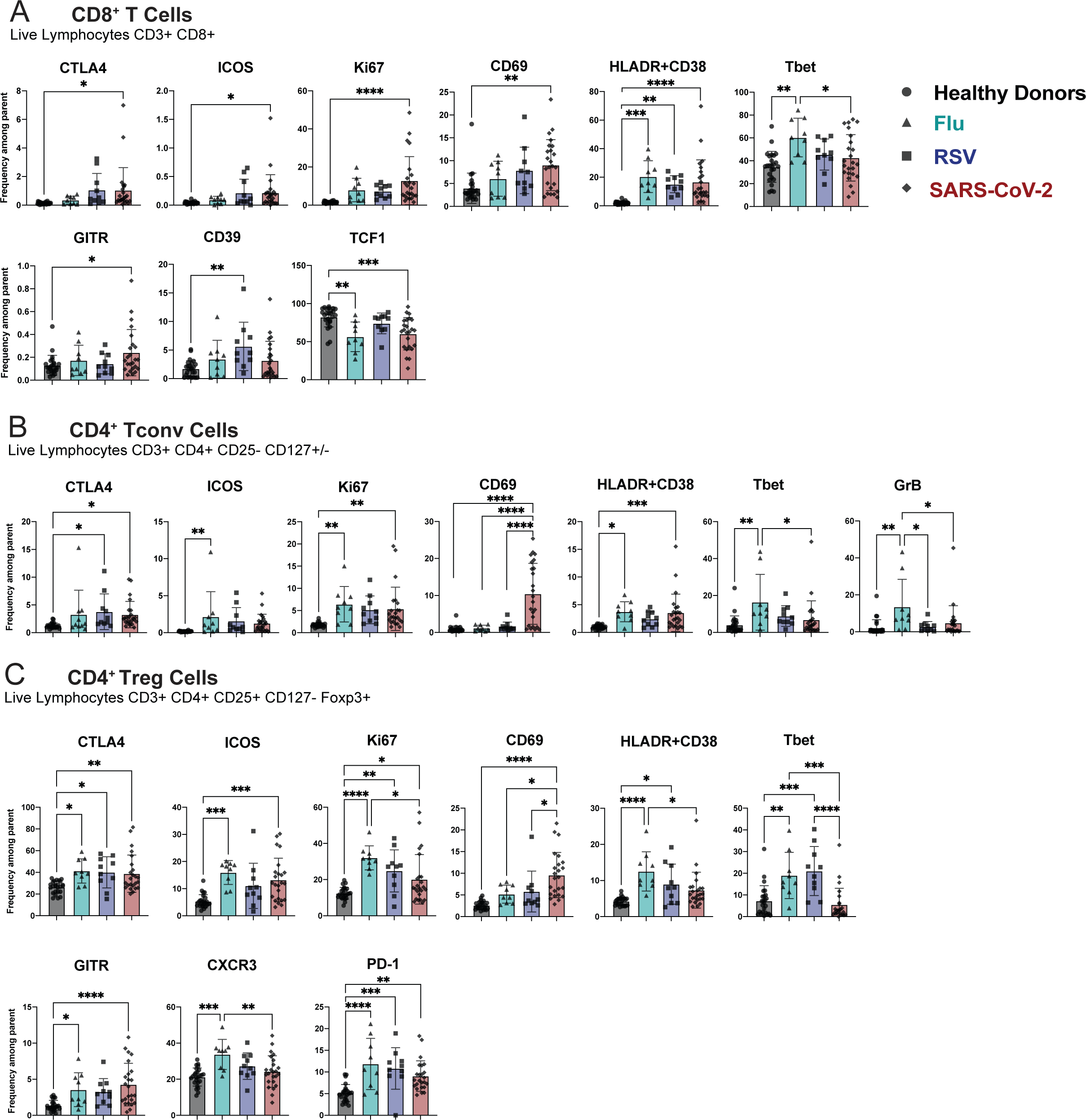
Phenotypical changes in T cell populations across respiratory infections. Bar graphs showing the mean frequency of indicated markers within the **A.** the CD8^+^ T cells, **B.** the CD4^+^ Tconv cells, **C.** the CD4^+^ Treg cells. Gating strategy of the different subsets can be found in Supplemental Figure 1. All data include at least 9 patients per group (Table 1) and are represented as mean ± SD. Depending on the distribution of our data, statistical analyses were performed using either one-way ANOVA or Kruskal Wallis test. * P < 0.05; ** P < 0.01; *** P < 0.001, **** P <0.0001.

**Supplemental Figure 5.**
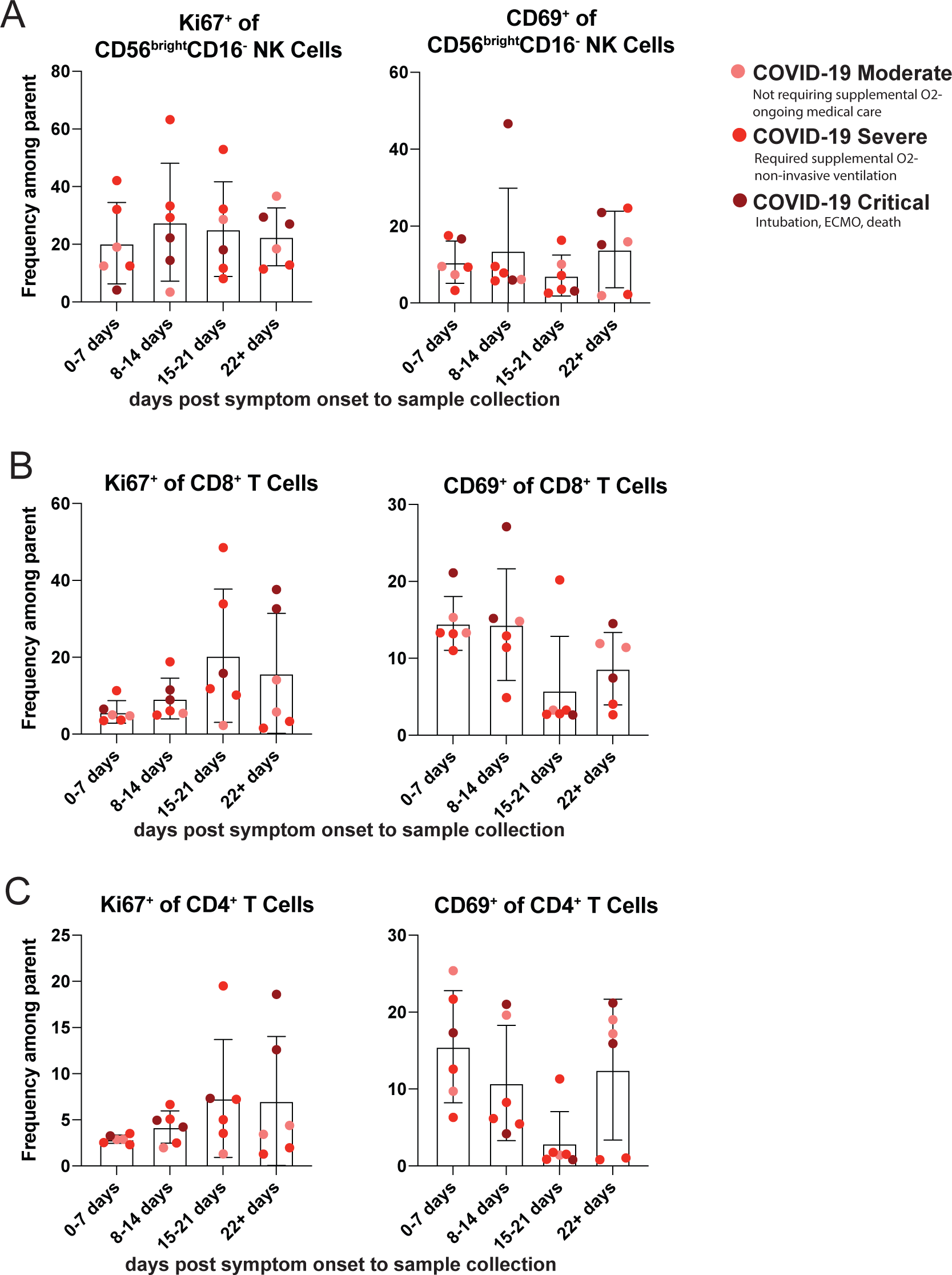
Markers of activation on NK and T cells by days post-symptom onset to sample collection. SARS-CoV-2 infected patients were grouped based on the reported days post-symptom onset to sample collection into four groups: 0-7 days, 8-14 days, 15-21 days, and 22 or greater days. The severity of COVID-19 disease is indicated by color of symbol. Markers of cellular activation were analyzed among **A.** CD56^bright^ CD16^-^ NK cells **B.** CD8^+^ T cells and **C.** CD4^+^T cells, for moderate COVID-19 (n=6), severe COVID-19 (n=12) and critical COVID-19 (n=6). All data are represented as mean ± SD. Depending on the distribution of our data, statistical analyses were performed using either one-way ANOVA or Kruskal Wallis test. * P < 0.05; ** P < 0.01; *** P < 0.001, **** P <0.0001.

